# Efficacy and safety of Mojeaga remedy in combination with conventional oral iron therapy for correcting anemia in obstetric population: a phase II randomized pilot clinical trial

**DOI:** 10.1101/2022.09.21.22280196

**Authors:** George Uchenna Eleje, Ifeanyichukwu Uzoma Ezebialu, Joseph Tochukwu Enebe, Nnanyelugo Chima Ezeora, Emmanuel Onyebuchi Ugwu, Iffiyeosuo Dennis Ake, Ekeuda Uchenna Nwankwo, Perpetua Kelechi Enyinna, Chukwuemeka Chukwubuikem Okoro, Chika Prince Asuoha, Charlotte Blanche Oguejiofor, Ejeatuluchukwu Obi, Chigozie Geoffrey Okafor, Angela Ogechukwu Ugwu, Lydia Ijeoma Eleje, Divinefavour Echezona Malachy, Chukwunonso Emmanuel Ubammadu, Emeka Philip Igbodike, Chidebe Christian Anikwe, Ifeoma Clara Ajuba, Chinyelu Uchenna Ufoaroh, Richard Obinwanne Egeonu, Lazarus Ugochukwu Okafor, Chukwunonso Isaiah Enechukwu, Sussan Ifeyinwa Nweje, Onyedika Promise Anaedu, Odigonma Zinobia Ikpeze, Boniface Chukwuneme Okpala, Ekene Agatha Emeka, Chijioke Stanley Nzeukwu, Ifeanyi Chibueze Aniedu, Emmanuel Chidi Chukwuka, Arinze Anthony Onwuegbuna, David Chibuike Ikwuka, Chisom God’Swill Chigbo, Chiemezie Mac-Kingsley Agbanu, Chidinma Ifechi Onwuka, Malarchy Ekwunife Nwankwo, Henry Chinedu Nneji, Kosisochukwu Amarachukwu Onyeukwu, Boniface Uwaezuoke Odugu, Sylvester Onuegbunam Nweze, Kenneth Chukwudi Eze, Shirley Nneka Chukwurah, Joseph Odirichukwu Ugboaja, Joseph Ifeanyichukwu Ikechebelu

## Abstract

**Background:** To our knowledge, there is no prior randomized trial on the effectiveness of Mojeaga remedy (a special blend of *Alchornea, Pennisetum, and Sorghum extracts*) when co-administered with standard-of-care for correction of anemia in obstetrics practice. This study determined the efficacy, safety and tolerability of Mojeaga as adjunct to conventional oral iron therapy for correction of anemia in obstetric population.

**Methods:** A pilot open-label randomized clinical trial. Participants with confirmed diagnosis of anemia in three tertiary hospitals in Nigeria were studied. Eligible participants were randomized 1:1 to either Mojeaga syrups 50 mls (200mg/50mls) administered three times daily in conjunction with conventional iron therapy (Mojeaga group) for 2 weeks or conventional iron therapy alone without Mojeaga (standard-of-care group) for 2 weeks. Repeat hematocrit level were done 2 weeks post-initial therapy. Primary outcome measures were changes in hematocrit level and mean hematocrit level at two weeks post therapy. Maternal adverse events and neonatal outcomes (birth anomalies, low birthweight, preterm rupture of membranes and preterm labor) were considered the safety outcome measures. Analysis was by intention-to-treat.

**Results:** Ninety five participants were enrolled and randomly assigned to the Mojeaga group (n=48) or standard-of-care group (n=47). The baseline socio-demographic and clinical characteristics of the study participants were similar. At two weeks follow-up the mean rise in hematocrit values from baseline (10.42±4.13% vs 6.36±3.69%;p<0.001) and mean hematocrit values (31.21±2.52% vs 27.7±3.49%;p<0.001) were significantly higher in the Mojeaga group. There were no treatment-related serious adverse events, congenital anomalies or deaths in the Mojeaga group and incidence of other neonatal outcomes were similar (p>0.05).

**Conclusion:** Mojeaga represents a new adjuvants for standard-of-care option for patients with anemia. Mojeaga remedy is safe for treating anemia during pregnancy and puerperium without increasing the incidence of congenital anomalies, or adverse neonatal outcomes. Clinical Trial Registration: **www.pactr.samrc.ac.za:** PACTR201901852059636 (https://pactr.samrc.ac.za/TrialDisplay.aspx?TrialID=5822).

## INTRODUCTION

Anemia in women is a major public health burden worldwide, particularly in low- and middle-income countries [1]. According to the World Health Organization (WHO), anemia affects approximately 2 billion people worldwide, which results in an estimated global prevalence of almost 25% [2]. The WHO estimates the prevalence of anemia among pregnant women to vary from 53.8% to 90.2% in low and middle-income countries, and 8.3–23% in high-income countries. The World Health Assembly has set a target of a 50% reduction in anemia among women of reproductive age by 2025 [1]. Although several factors have been implicated, iron deficiency is undoubtedly the most common cause of anemia worldwide [3, 4, 5].

Iron deficiency anemia (IDA) is often treated with oral iron supplement even in pregnancy. However, due to gastrointestinal side effects such as nausea, vomiting, and constipation, compliance is often poor and results in subsequent discontinuation [4, 6, 7]. As such, intravenous iron administration or blood transfusion is increasingly being recommended for women who are non-compliant with oral iron, have severe IDA, or who require rapid intervention [6, 7]. However, intravenous drug administration has been infrequently utilized in clinical practice due to undesirable adverse reactions, including severe allergic reactions and anaphylaxis [8]. Blood transfusion is not without complications and some religious groups object to blood transfusions all over the world including Nigeria. Hence, the need to sort for a safer and self-administered option to address the menace of IDA especially in low and middle-income countries.

Mojeaga herbal remedy (produced by Mojeaga International Ventures Ltd, Nigeria) is a natural preparation containing a combined *Alchornea, Pennisetum*, and *Sorghum* extracts [8, 9]. It contains free organic oral iron preparation among other nutrients that allows co-administration of conventional hematinics or other iron preparations. Mojeaga has been approved under Listing status by the National Agency for Food and Drug Administration and Control (NAFDAC) with NAFDAC registration number of A7-0996L as safe for use while allowing data generation on efficacy through clinical trial, for which this research stood in the pathway of providing scientific evidence for safety and efficacy. The *Pennisetum* extract and *Sorghum* extract in Mojeaga are excellent sources of iron, potassium, flavonoids and other phytonutrients. According to the manufacturers, Mojeaga works by promoting healthy metabolic processes because it contains high levels of B-group vitamins [8, 9]. Mojeaga also works by effectively mopping up the free radicals and reactive oxygen species, and quickly reverse the lipid peroxidative and cellular damages. It increases the levels of packed cell volume and hemoglobin and contains antioxidants and natural minerals. It also has strong anti-inflammatory properties and enhances general body metabolism. It builds the immune system, and it is high in natural iron [8, 9]. There is an additional major benefit of the Mojeaga, in that it can be administered in relatively high normal doses in a short period of time [10].

In one recent report, anemic pregnant women of gestational age of 36 weeks with IDA of hemoglobin level of 5g/dl received Mojeaga in addition to oral iron therapy (adjunct therapy), and had their hemoglobin appreciated to 10 g/dl two weeks after treatment without any adverse pregnancy or neonatal outcomes [8]. In other published reports, a significantly higher number of women achieved anemia correction within a shorter time frame, and there were markedly fewer gastrointestinal treatment-related adverse events especially when the drug was administered in sips [9, 10, 11]. This lends support to the hypothesis that Mojeaga in combination with conventional oral iron therapy is superior to conventional oral iron therapy alone for correcting anemia in the obstetric population.

To the best of our knowledge, this is the first randomized trial on the efficacy and safety of combined Mojeaga and oral iron therapy schedule. Therefore, this study determined the effectiveness, safety and tolerability of Mojeaga as an adjunct to conventional oral iron therapy (standard-of-care) for correcting anemia in obstetric population.

## METHODS

### Study setting

The study was carried out at the antenatal (obstetrics) clinics of Nnamdi Azikiwe University Teaching Hospital (NAUTH), Nnewi, Enugu State University of Science and Technology Teaching hospital, Parklane, Enugu and Chukwuemeka Odumegwu Ojukwu University Teaching Hospital, Awka, all are tertiary hospitals in the South-east Nigeria.

### Study design

This was an open-label randomized clinical trial.

### Study population

The participants comprised of obstetrics participants with clinical and laboratory diagnosis of anemia and gave written informed consent before recruitment. The participants were recruited at the antenatal clinics or referred for treatment of their anemia from other hospitals. Participants were recruited prior to commencing the intervention and control agents.

### Inclusion criteria

Inclusion criteria included obstetrics adult participants with confirmed clinical and laboratory diagnosis of anemia (hematocrit of <31.5%) and with normal liver and renal function markers or profiles. Anemia was confirmed by prospective expert hematology review. In this study, significant anemia in pregnancy is defined as a hemoglobin concentration <11 g/dL (or hematocrit <33.0%) in the first trimester or <10.5 g/dL (or hematocrit <31.5%) in the second and third trimesters [12].

### Exclusion criteria

Pregnant women in the first trimester of pregnancy (because of nausea and vomiting during this period), chronic medical disorders including HIV/AIDS, cancers, etc. and women on chronic medications that cause anemia were excluded.

### Randomization and Allocation Sequence

Following consent, patients at the selected hospitals were screened for eligibility. The particpants, eligible for the study, were randomized into two groups (1:1 ratio, blocks of 4) using simple (block) randomization using a randomization table created by a computer software program by a person not involved in the study and available at https://mahmoodsaghaei.tripod.com/Softwares/randalloc.html. Allocation sequences and codes were concealed from the person allocating the participants to the intervention arms using numbered containers containing the drugs.

### Blinding of participants, Personnel and Outcome assessors

Only the outcome assessors were blinded. The study was open-label, with both participants and investigators aware of treatment assignment.

### Study Procedure/Drug Administration

Participants with a clinical and laboratory diagnosis of anemia presenting in Outpatient Clinic or antenatal clinic of the study hospitals for symptoms or signs of anemia were screened consecutively. All participants underwent routine medical examination that included pulse rate, body weight, blood pressure determination and general examination to ascertain the presence of and severity of anemia. All consenting participants were diagnosed to have either anemia or not after undergoing the hematological test. Only patients with confirmed clinical and laboratory diagnosis of anemia were randomized. Eligible participants were sequentially allocated using an opaque sealed envelope to receive either Mojeaga and conventional oral Therapy, or the conventional oral iron Therapy alone. Therapies were given for two weeks.

### Intervention Therapy

Standard doses of 50 mls (200mg/50mls) of Mojeaga were administered three times daily in conjunction with conventional iron therapy (standard-of-care) two times a day (breakfast and dinner) for 2 weeks.

### Control Therapy

Standard/conventional doses of iron therapy (standard-of-care) were administered two times a day (breakfast and dinner) for 2 weeks. These control participants with confirmed diagnosis of anemia were placed on conventional iron therapy administered without Mojeaga.

### Follow-Up

The trial lasted for 12 months. Recruitment was within the first 6 months of commencement of the study and the follow-up was for at least 6 months from recruitment. All participants were followed up in outpatient settings. During each follow-up weekly visit, participants were contacted on phone on daily basis to assess the level of compliance on the trial drugs. Participants were informed about the usual side effect of hematinic preparations and were told to report nausea, vomiting, bowel disturbances, or any other complications. Where possible, the participants were also encouraged to record any side effects or adverse events in a paper that was reviewed at each follow-up visit, and they were explicitly asked about such events during each interview. Where possible, the drug compliance were checked before follow-up test and at follow-up visit by checking the used drug packets. Any participants found to be developing complications such as worsening of the symptoms from the study were given appropriate treatment. Repeat hematocrit level was done 2 weeks post initiation of treatment in all the participants to confirm or refute success of the treatment (correction of anemia and levels of hematocrit) and absence/presence of adverse effects. All the pre and post (repeat) hematocrit level were carried out by a Hematologist, while the liver function test (LFT) and serum electrolytes, urea and creatinine (SEUCR) were carried out by the senior laboratory Scientist of Chemical Pathology in Nnamdi Azikiwe University Teaching Hospital, Nnewi, Nigeria laboratory and in the other two collaborating hospitals.

## OUTCOME MEASURES

### Primary

The primary endpoint included changes in the hematocrit level and mean hematocrit level at two weeks after initial therapy.

### Secondary

The secondary endpoints included the proportion of patients with persisting anemic symptoms (epigatric pain, weakness, dizziness) at two weeks after initial therapy, incidence of any maternal adverse events (such as diarrhea, nausea, vomiting, colitis and drop-out from adverse effects) following commencement of therapy, incidence of any fetal adverse events (such as preterm labor, preterm premature rupture of membranes, low birthweight or birth anomaly) following commencement of therapy, mean levels of renal function parameters (serum electrolyte, urea and creatinine) at two weeks after initial therapy and mean levels of liver function parameters (aspartase transaminase (AST); alkaline phosphatase (ALP); and alanine transaminase (ALT)) at two weeks after initial therapy. As safety variables, adverse events were monitored throughout the whole study period from the signature of the informed consent form up to the last visit.

### Sample size determination

Allowing for 20% attrition, a sample size of 47 participants per group yields 90% power to detect a difference of 3 percent on the original hematocrit level [8] ranging from 27 to 30 using a two-sample means test at a two-sided alpha of 0.05, assuming a standard deviation of 2.9.

### Sampling Approach

Simple random sampling method was used in which consecutive, eligible, consenting women with diagnosis of anemia were randomly allotted into two equal groups of using a random number table.

### Drug active dose validation/standardization

The cover of the Mojeaga remedy bottle has a measuring cap which is of 25 mls capacity for uniform administration and validation. Each 500mls of Mojeaga remedy contains 2069.57 mg of active Mojeaga remedy [13, 14]. Therefore, each 50mls of Mojeaga contains approximately 200mg of active Mojeaga remedy. Therefore the dose of Mojeaga remedy of 200mg/50mls three times daily for two weeks was utilized.

### Statistical Analysis

Data was analyzed using SPSS version 23, IBM Company, USA. The data were expressed as the number (percentage), mean (standard deviation [SD]), and mean (95% confidence interval [95% CI] as appropriate. Categorical variables were compared using the Chi-squared test and Fisher’s exact test, as necessary and relationship expressed using relative risks and 95% confidence intervals. Independent t-test or Mann-Whitney U-test was used to compare mean of continuous variables between treatment groups, depending on their normality of distribution. The intention-to-treat efficacy analyses was based on all the patients who received the study medication and had completed the follow-up visit. Interim analyses were done after 30 participants have been recruited. Participants with no observed outcome were considered as treatment failures. When possible, subgroup analysis were also done, where applicable. A p value of <0.05 was considered to be significant. Interim analyses of principal safety and effectiveness outcomes were performed on behalf of the data and safety monitoring committee by the trial statistician (who remained unaware of the treatment assignments).

### Ethical consideration

The study adhered to CONSORT guidelines [15]. The study protocol was approved by the Nnamdi Azikiwe University Teaching Hospital, Nnewi, Nigeria ethics committee and other collaborating hospitals, with the approval numbers: NAUTH/CS/66/VOL.12/014/2019/008, ESUTHP/C-MAC/RA/034/103, and COOUTH/CMAC/ETH.C/VOL.1/0056. The study was registered and approved by the Pan African Clinical trial registry: at https://pactr.samrc.ac.za/, with approval number of **PACTR201901852059636**. The procedures followed was in accordance with the guidelines of the World Medical Association’s Declaration of Helsinki (1964, and its later amendments). The drug (Mojeaga) was registered with NAFDAC Nigeria with registration number of NRN: A7-0996L. The trial was also registered and approved by National agency for food drug and control (NAFDAC) with NAFDAC Trial Registration No of NAF/DER/LAG/V&CT/MOJEAGA/2021. The nature of the study were carefully explained to the participants and their written consent obtained before being recruited into the study. The rights of the participants to participate or withdraw from the study were fully honored without any adverse consequence to the participants. All trial participants were provided life insurance throughout the duration of the study.

### Certification of Analysis

To ensure a high quality standardized formulations, the raw material was authenticated and the product was laboratory tested and certified by Prof JU Iyasele of Chemistry Department, University of Benin, Nigeria in accordance with Institute of Public analyst of Nigeria Decree no 100 of 1992.

## RESULTS

### Baseline characteristics

Between February 27, 2020, and February 20, 2022, two hundred and sixty seven participants were assessed for eligibility, 72 participants were excluded for different reasons (58 did not meet the inclusion criteria: HIV/AIDS (25); first trimester pregnancy (17) and declined to participate (16)), while 95 eligible participants were enrolled and randomly assigned to the Mojeaga group (n=48) or the standard-of-care group (n=47; figure 1).

**Figure 1:**
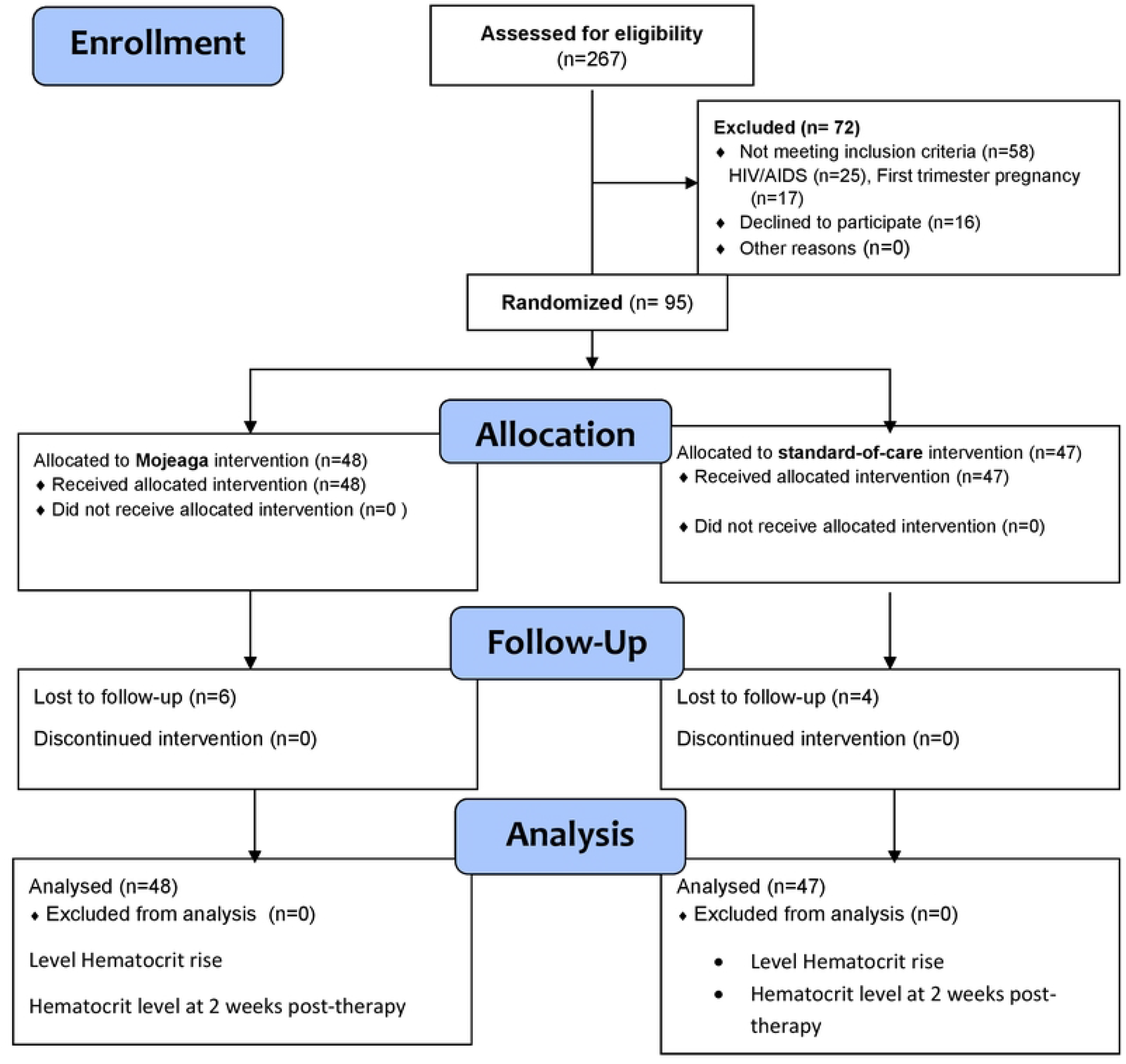
Flow chat of the participants.

Of the 95 participants, 85 were pregnant: Mojeaga group (n=42) and standard-of-care group (n=43), while 10 were puerperal women (Mojeaga group (n=6) and standard-of-care group (n=4). So far, all the 95 participants completed the treatment and were included in analysis.

As shown in Table 1, the baseline socio-demographic and clinical characteristics of the study participants including age, gestational age at recruitment, body mass index, mean duration of treatment, e.t.c., were not significantly different between the 2 groups (p> 0.05). The details are as shown in Table 1.

**Table 1:**
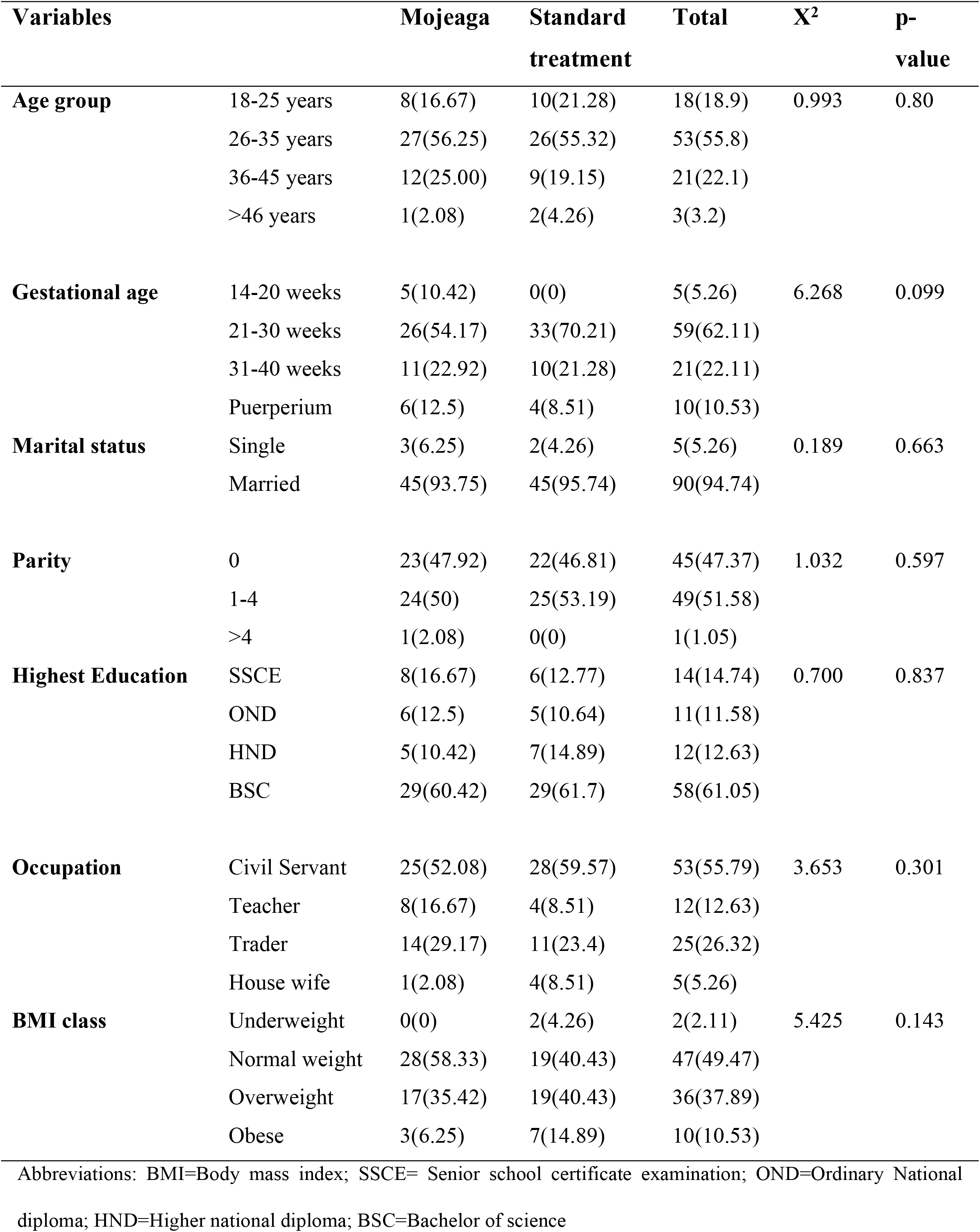
Distribution of sociodemographic variables across research groups.

Additionally, the baseline pre-therapy hematocrit level, pre-therapy serum electrolyte, urea and creatinine level and liver function test parameters were similar in both groups (p>0.05). The details are as shown in table 2.

**Table 2:** Comparison of pre-treatment serum variables between research groups

| **Research Groups** | **Mojeaga**<br>**Mean±STD** | **Standard**<br>**Mean±STD** | **Mojeaga**<br>**Rank** | **Standard**<br>**Rank** | **U-value** | **p-value** |
| --- | --- | --- | --- | --- | --- | --- |
| **pre-Na (mmol/L)** | 136.45±1.89 | 137.76±4.29 | 43.64 | 52.46 | 918.50 | 0.12 |
| **pre-K (mmol/L)** | 3.38±0.24 | 3.39±0.32 | 51.17 | 44.77 | 976.00 | 0.25 |
| **pre-Cl (mmol/L)** | 100.44±2.93 | 99.38±0.97 | 52.78 | 43.12 | 898.50 | 0.08 |
| **pre-Bicarbonate**<br>**(mmol/L)** | 25.83±0.7 | 25.94±1.65 | 43.07 | 53.03 | 891.50 | 0.07 |
| **pre-Urea (mmol/L)** | 2.45±0.18 | 2.42±0.26 | 49.14 | 46.84 | 1073.50 | 0.67 |
| **pre-Creatinine (μ/L)** | 95.32±6.25 | 95.7±6.3 | 43.02 | 53.09 | 889.00 | 0.07 |
| **pre-AST (μ/L)** | 12.18±5.39 | 11.89±3.83 | 43.80 | 52.29 | 926.50 | 0.12 |
| **pre-ALP (μ/L)** | 61.55±8.79 | 60.45±16.33 | 47.15 | 48.87 | 1087.00 | 0.76 |
| **pre-ALT (μ/L)** | 10.88±4.05 | 10.17±2.79 | 49.11 | 46.86 | 1074.50 | 0.69 |
| **pre-hematocrit**<br>**(%)** | 20.79±3.91 | 21.34±3.58 | 46.01 | 50.03 | 1032.5 | 0.47 |

Abbreviations: Na (NR) =Sodium (Normal range: 135-145); K=Potassium (NR 3.5-5.5); Cl=Chloride (NR 96-106); Bicarbonate (NR 21-31); Urea (NR 1.7-9.1); Creatinine (NR 53-106); AST= Aspartase transaminase (NR 1-40); ALP= Alkaline phosphatase (NR 60-170); ALT= Alanine transaminase (NR 1-40); Hematocrit (NR 30-54)

### Primary endpoints

#### Mean rise in hematocrit values

The mean rise in hematocrit values at two weeks follow-up was significantly higher in Mojeaga group (10.42±4.13% vs 6.36±3.69%; p<0.001). Similarly, the mean hematocrit values at two weeks follow-up was significantly higher in Mojeaga group (31.21±2.52% vs 27.7±3.49%; p<0.001). This is shown in table 3.

**Table 3:** Comparison of haematocrit parameters (primary outcome) among research groups after treatment.

|  |  | **Mean Rank** | **Mean** | **U-value** | **p-value** |
| --- | --- | --- | --- | --- | --- |
| **Haematocrit rise** | Mojeaga (%) | 61.48 | 10.42±4.13 | 481 | <0.01 |
|  | Standard treatment (%) | 34.23 | 6.36±3.69 |  |  |
| **post-haematocrit** | Mojeaga (%) | 63.03 | 31.21±2.52 | 406 | <0.01 |
|  | Standard treatment (%) | 32.65 | 27.7±3.49 |  |  |

### Secondary endpoints

Table 4 shows the comparison of symptom resolution between the two research groups pre and post-therapy. There is no difference in tolerability and outcomes in other clinical parameters between the two groups (p>0.05). Mojeaga was associated with greater improvement in dizziness and weakness scores. No serious adverse events were noted in either group. There was no recorded maternal or fetal death.

**Table 4:** Comparison of symptom resolution between research groups pre and post-therapy

|  |  | **Mojeaga** | **Standard** | **X<sup>2</sup>** | **p-value** |
| --- | --- | --- | --- | --- | --- |
| **pre-Epigastria pain** | Present | 8(16.67) | 6(12.77) | 0.773 | 0.403 |
|  | Absent | 40(83.33) | 41(87.23) |  |  |
| **pre-Dizziness** | Present | 20(41.67) | 22(46.81) | 0.682 | 0.383 |
|  | Absent | 28(58.33) | 25(53.19) |  |  |
| **pre-Weakness** | Present | 28(58.33) | 25(53.19) | 0.682 | 0.383 |
|  | Absent | 20(41.67) | 22(46.81) |  |  |
| **pre-Hematemesis** | Present | 0(0) | 0(0) | - | - |
|  | Absent | 48(100) | 47(100) |  |  |
| **post-Epigastria pain** | Present | 0(0) | 0(0) | - | - |
|  | Absent | 48(100) | 47(100) |  |  |
| **post-Dizziness** | Present | 0(0) | 0(0) | - | - |
|  | Absent | 48(100) | 47(100) |  |  |
| **post-Weakness** | Present | 2(4.17) | 4(8.51) | 0.435 | 0.329 |
|  | Absent | 46(95.83) | 43(91.49) |  |  |
| **post-Hematemesis** | Present | 0(0) | 0(0) | - | - |
|  | Absent | 48(100) | 47(100) |  |  |

### Proportion of participants with persisting anemic symptoms (epigatric pain, weakness, dizziness) at two weeks after initial therapy

Persistent anemic symptoms were reported in 4.2% of the Mojeaga group and 8.5% of the control group (p = 0.329). Figure 2 shows the presence of symptoms before and after treatment in the both groups.

**Figure 2:**
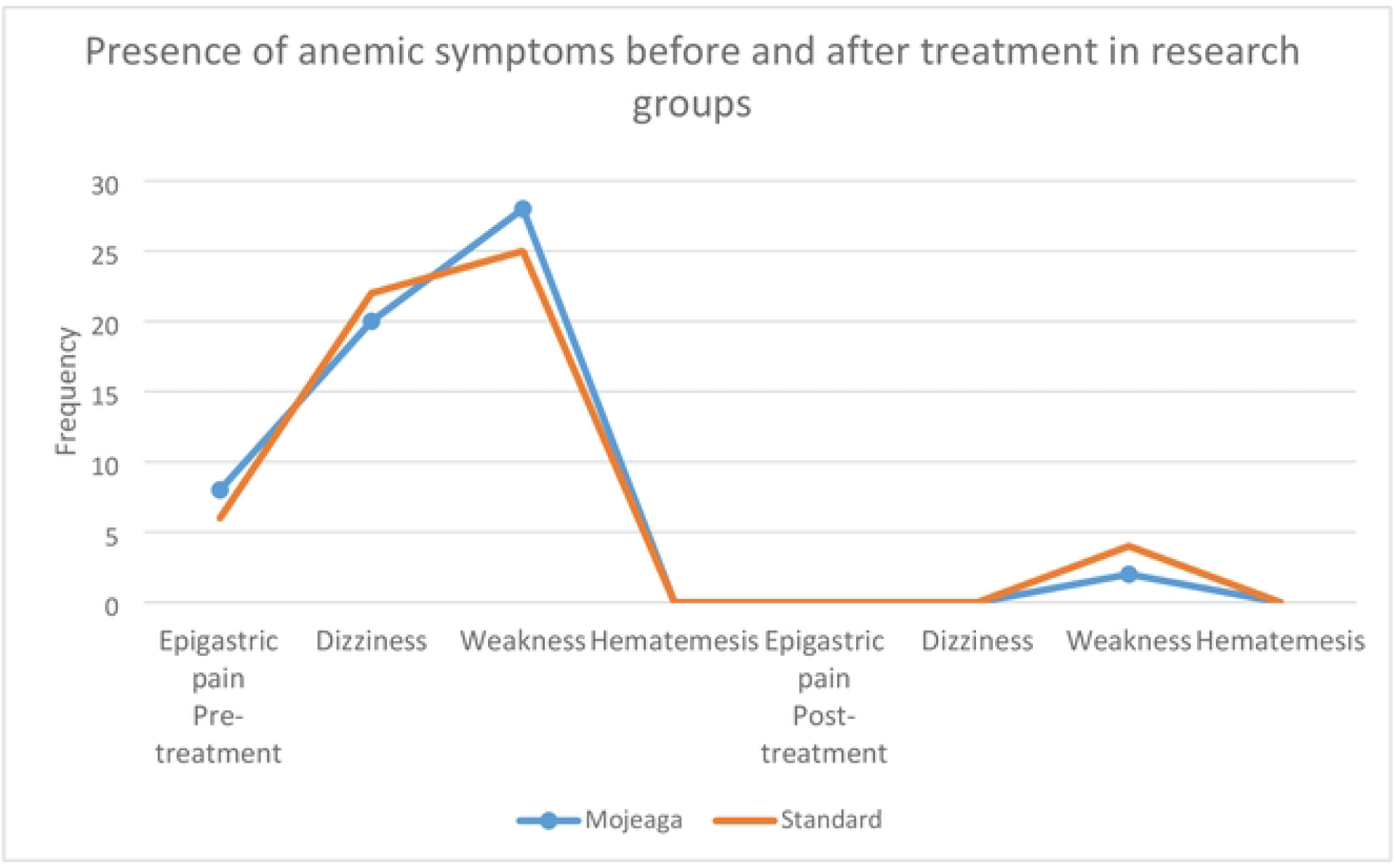
Presence of symptoms before and after treatment in the Mojeaga group and Standard-of-care group.

### Incidence of fetal adverse events (such as congenital anomaly, preterm labor, PROM or low birth weight) following commencement of therapy

Table 5 shows the comparison of incidence of fetal adverse events between the two groups. Both groups experienced comparable decrease in fetal adverse events. There were no recorded congenital anomaly and no changes in incidence of preterm labor, and low birth weight in either group.

**Table 5:** Comparison of incidence of fetal adverse events among research groups.

|  |  | **Mojeaga** | **Standard treatment** | **p-value** |  |
| --- | --- | --- | --- | --- | --- |
| **Preterm Labour** | Yes | 2(4.17) | 2(4.26) | 0.983 |  |
|  | No | 46(95.83) | 45(95.74) |  |  |
| **Preterm PROM** | Yes | 0(0.00) | 2(4.26) | 0.149 |  |
|  | No | 48(100.00) | 45(95.74) |  |  |
| **Birth weight class** | Not available | 6(12.50) | 4(8.51) | 0.237 |  |
|  | Normal birth weight | 39(81.25) | 35(74.47) |  |  |
|  | Low birth weight | 3(6.25) | 8(17.02) |  |  |
| **Birth anomalies** | Yes | 0(0.00) | 0(0.00) | - | - |
|  | No | 48(100.00) | 47(100.00) |  |  |

Abbreviations: PROM=Premature rupture of fetal membranes

### Incidence of maternal adverse events (such as diarrhea, nausea, vomiting, colitis and drop out due to adverse effects) following commencement of therapy

Table 6 shows the prevalence of adverse effects in research groups during treatment. Figure 3 shows the presence of adverse effects during and after treatment in the two groups. Both groups experienced comparable low numbers of maternal adverse events. Maternal adverse events were reported in 6.25% of the Mojeaga group and 10.64% of the control group but the difference was not significant (p=0.19).

**Table 6:** Frequency of adverse effects in research groups during treatment

|  | **Mojeaga** | **Standard** | **X<sup>2</sup>** | **p-value** |
| --- | --- | --- | --- | --- |
| **Nausea** | 3(6.25) | 5(10.64) | 6.64 | 0.19 |
| **Vomiting** | 0(0) | 0(0) | - | - |
| **Colitis** | 0(0) | 0(0) | - | - |
| **Drop-out from adverse effect** | 0(0) | 0(0) | - | - |

**Figure 3:**
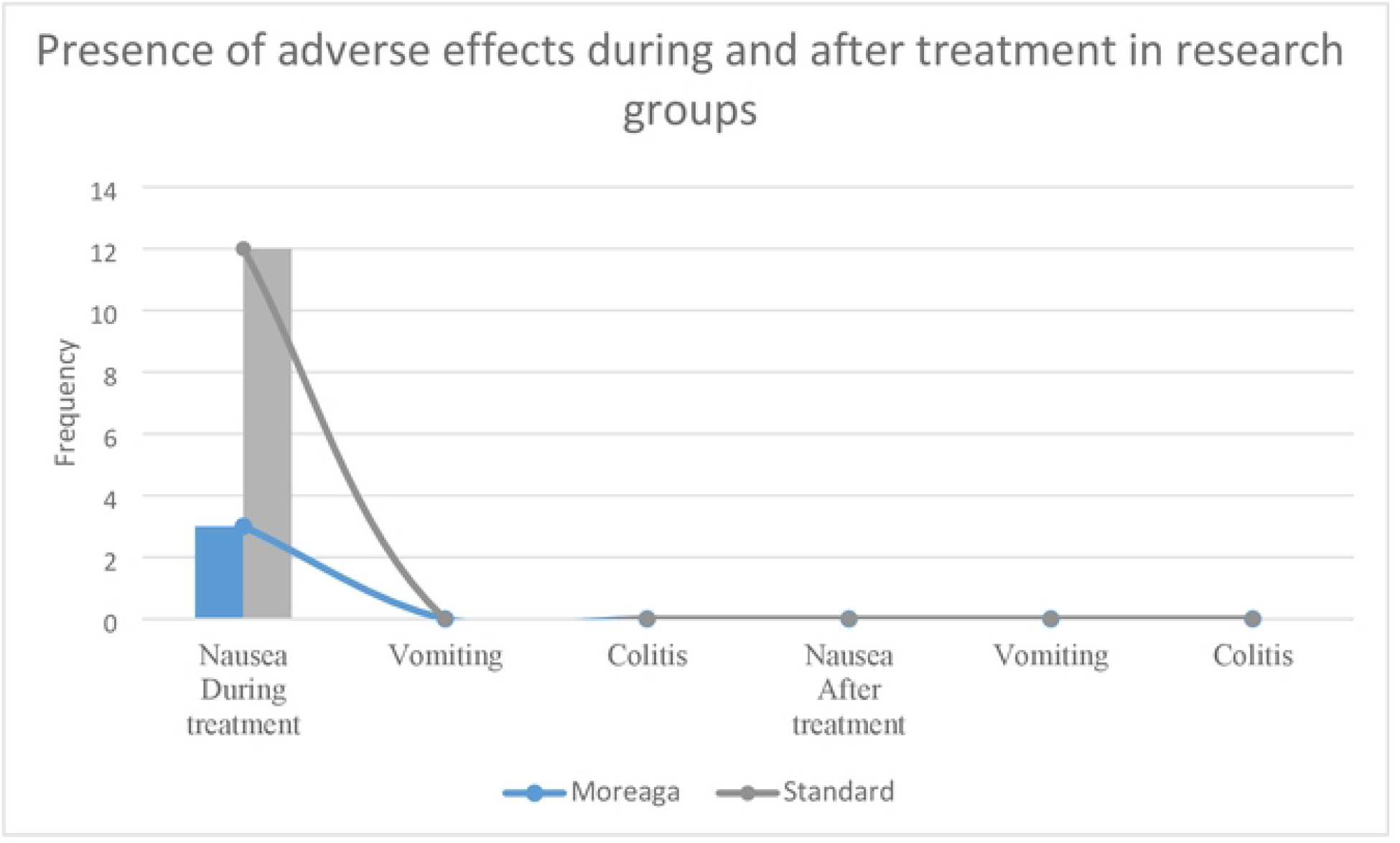
Presence of adverse effects during and after treatment in the two groups

### Mean levels of renal function parameters (serum electrolyte, urea and creatinine) at two weeks after initial therapy

Table 7 shows the comparison of serum electrolyte, urea and creatinine as well as liver function tests between the two groups at post-therapy. Serum bicarbonate level was significantly lower in Mojeaga group than control group (22.49±2.07 vs 24.02±2.09, P = 0.002). Other electrolytes, and urea and creatinine levels were similar in both groups (p > 0.05).

**Table 7:** Comparison of serum electrolyte and function tests of all research participants at post-therapy.

|  | **Research Groups** | **Rank** | **Mean±SD** | **U-value** | **p-value** |
| --- | --- | --- | --- | --- | --- |
| **post-Na** | Mojeaga | 44.74 | 136.06±3.54 | 972 | 0.239 |
|  | Standard treatment | 51.33 | 136.62±3.64 |  |  |
| **post-K** | Mojeaga | 47.55 | 3.47±0.28 | 1107 | 0.867 |
|  | Standard treatment | 48.46 | 3.51±0.36 |  |  |
| **post-Cl** | Mojeaga | 52.07 | 101.04±4.38 | 933 | 0.141 |
|  | Standard treatment | 43.84 | 99.9±3.95 |  |  |
| **post-Bicarbonate** | Mojeaga | 39.86 | 22.49±2.07 | 738 | 0.002 |
|  | Standard treatment | 56.31 | 24.02±2.09 |  |  |
| **post-Urea** | Mojeaga | 43.64 | 2.26±0.56 | 919 | 0.110 |
|  | Standard treatment | 52.46 | 2.27±0.51 |  |  |
| **post-Creatinine** | Mojeaga | 51.73 | 86.02±10.14 | 949 | 0.162 |
|  | Standard treatment | 44.19 | 86.97±8.37 |  |  |
| **post-AST** | Mojeaga | 50.28 | 17.5±7.91 | 1019 | 0.409 |
|  | Standard treatment | 45.67 | 15.2±7.59 |  |  |
| **post-ALP** | Mojeaga | 51.28 | 66.99±19.65 | 971 | 0.236 |
|  | Standard treatment | 44.65 | 64.01±15.85 |  |  |
| **post-ALT** | Mojeaga | 43.79 | 8.62±4.41 | 926 | 0.122 |
|  | Standard treatment | 52.3 | 8.43±2.38 |  |  |

Abbreviations: Na (NR) =Sodium (Normal range: 135-145); K=Potassium (NR 3.5-5.5); Cl=Chloride (NR 96-106); Bicarbonate (NR 21-31); Urea (NR 1.7-9.1); Creatinine (NR 53-106); AST= Aspartase transaminase (NR 1-40); ALP= Alkaline phosphatase (NR 60-170); ALT= Alanine transaminase (NR 1-40); Hematocrit (NR 30-54)

### Mean levels of liver function parameters (AST, ALT and ALP) at two weeks after initial therapy

As shown in Table 6, there were no significant changes in liver function parameters including AST, ALT and ALP in the two groups.

## DISCUSSION

This trial which is the first positive randomized trial involving Mojeaga therapy in women with anemia shows that Mojeaga therapy improves anemia compared with the use of standard of care alone. Until now, treatment recommendations for moderate and severe anemia in pregnancy and gynecological patients were based on the use of parenteral therapy and blood transfusion and occasionally oral therapy. The findings of this study suggest that concomitant use of standard iron therapy with Mojeaga improves efficacy with significant rise in hematocrit and higher hematocrit level.

In this study, the mean rise in hematocrit values at two weeks follow-up was significantly higher in Mojeaga group compared to the standard-of-care group (10.42±4.13% vs 6.36±3.69%; p<0.001). Also, the mean hematocrit values at two weeks follow-up was significantly higher in Mojeaga group (p<0.001). The apparent anemia recovery benefit of Mojeaga regardless of pregnant or no-pregnant status suggests that Mojeaga is promising adjuvant therapy for anemia. This benefit could be due to high amount of natural iron in the Mojeaga formulations. Similarly, the previous case report by Eleje et al on the use of Mojeaga re-counted improvements in anemia symptoms in women that received Mojeaga remedy [8]. In another recent study by Idu et al aimed at evaluating the hematinic property of Mojeaga herbal remedy in animal model, [13], the hematological indexes showed a significant increase in red blood cells, and hematocrit at day 10 of the experimental rats with the values of red blood cells (6.72, 7.34, 7.10 × 10^6^/ul) and hematocrit (47.8, 51.5, 49.75%) respectively higher in Mojeaga group when compared with the control (*p* < 0.05). However, Idu et al study did not directly evaluate the Mojeaga in human subjects [13]. Similarly, in participants given Mojeaga in our study, improved return in anemia within 2 weeks of follow-up were both markedly higher in the participants that received Mojeaga than controls (31.21±2.52% vs 27.7±3.49%; p<0.001). This earlier return of normal blood levels will be beneficial for surgical patients since they will meet up with the pre-anesthetic requirement of normal hematocrit of >30%.

Nevertheless, although this emerging pilot data is of interest, until more definitive information regarding the association between Mojeaga intake and improvement of anemia becomes available, Mojeaga should be considered as an adjuvant option for all pregnant women with anemia. Mojeaga has shown an associated increase in the probability of response to standard of care, and showed a trend toward an overall improved benefit.

Iron deficiency anemia (IDA) during pregnancy is associated with an increased risk of preterm birth, low birthweight, fetal growth restriction, and increased neonatal and maternal mortality. Furthermore, iron deficiency may predispose a person to postpartum IDA, peripartum blood transfusion, infections, and precipitate heart failure [6].

The findings of our study raise the question of the mechanism of action of Mojeaga in the treatment of anemia in obstetrics population. It is possible that Mojeaga works by rapid turn-over of the bone-marrow erythropoiesis. This possible mechanisms was revealed by the findings of Idu et al that showed that the histo-architectural structure of the bone marrow of the experimental animals showed a stimulating effect of myeloid/erythroid cell ratio > 60 in the treatment (Mojeaga) groups when compared with control groups. In many institutions, first-line oral therapy is polymaltose, astyfer, and fesolate. In such participants, the results of our study would strongly suggest that Mojeaga would be the most favorable adjuvant option at first consideration.

Our randomized controlled clinical trial showed that Mojeaga has a favorable safety profile, and also when combined with the conventional oral iron therapy. The frequency of adverse events, abnormal laboratory analysis, vital signs, and physical findings was similar across the two treatment groups. Although serum bicarbonate level was different from both study groups, the overall bicarbonate levels were within the normal limits. Similarly, in a recent study by Idu et al that evaluated the toxicological profile of Mojeaga herbal remedy on male and female animal models, acute and chronic toxicity of Mojeaga herbal remedy in male and female Wistar rats were investigated through thorough examination of mortality rate, body and organ weight changes, hematological indexes, biomarkers of hepatic and renal functions, lipid profile, in-vivo antioxidant assay, hormonal assay and histopathological study across all treatment groups using standard protocol [14]. There was no observable behavioral change with absent lethality at 10 to 10000 mg/kg of Mojeaga remedy. There was also no drastic significant change (p>0.05) in the body and organ weight of the experimental animals. In the chronic toxicity arm of the study, Mojeaga remedy indicated no significant difference (p > 0.05) in hematological indices, liver function test, kidney function test, lipid profile, antioxidant indexes and hormonal assays with a slight significant increase (p < 0.05) in hepatic (ALT, ALP, AST) and renal (potassium, sodium and chloride), and lipid profile (cholesterol, triglyceride, high density lipoprotein, low density lipoprotein). There was no marked significant toxicological effect (p>0.05) on serum total protein, blood urea nitrogen, albumin, creatinine and urea levels across the whole treated groups at graded doses of mojeaga. Mojeaga product caused no histopathological variation on vital visceral organs (liver, kidney, heart, stomach, brain, lungs spleen testes and uterus) when compared with controls [14].

The strengths of the study include the good uptake and inclusion of a representative sample of participants from obstetrics units across a range of socioeconomic groups and hospitals, the near-complete follow-up period allowing complete case analysis, and limiting bias in the results. Therapeutic options for women with anemia especially those who refuse blood transfusion have been scarce, in part because recruitment challenges have made randomized controlled trials in cases with anemia requiring oral therapy infeasible. Our study has overcome this challenge. This study was the first randomized trial to evaluate the effectiveness, safety and tolerability of Mojeaga as adjunct to conventional oral iron therapy for correction of anemia in obstetrics population. One potential weakness of this study was the bias of individual investigators to open label nature of the trial. To control this, the protocol required blinded intervention sequence and computer-generated random numbers. Additionally, the intention-to-treat analysis was employed with resultant marked benefit associated with Mojeaga, supporting the evidence for improvement in anemia symptoms. While hemoglobin concentration, hematocrit, and to a lesser extent erythrocyte count, are the anemia indicators used in clinical practice, these parameters are only surrogates for the actual definition of anemia: a reduction in erythrocyte mass per unit body weight [16].

## CONCLUSION

In conclusion, based on the findings of our study, Mojeaga should be considered a new adjuvants to standard-of-care option for pregnant and puerperal women with anemia. Mojeaga remedy is safe for treating anemia during pregnancy and puerperium without increasing the incidence of congenital anomalies, low birthweight, preterm labor/rupture of membranes or other adverse events. These findings, therefore, suggest the potential benefit of Mojeaga for treating patients with anemia and support for further adequately powered confirmatory trials investigating the efficacy and safety of Mojeaga remedy.

## Data Availability

All relevant data are within the manuscript and its Supporting Information files.

## Acknowledgements

The study was coordinated by the Effective Care Research Unit at Nnamdi Azikiwe University University, Awka, Nigeria. The authors appreciate the help of the staff of Nnamdi Azikiwe University University Teaching Hospital (NAUTH), Nnewi, Nigeria; Enugu State University of Science and Technology Teaching hospital, Parklane, Enugu, Nigeria and Chukwuemeka Odumegwu Ojukwu University Teaching Hospital, Awka, Nigeria and participants involved in the trial. Publication of these results should not be considered an endorsement of any product used in this study by the Nnamdi Azikiwe University or any of the organizations where the authors are affiliated.

## Authors’ contributions

GUE, IUE and JTE designed the study and carried out the procedures for this project, while other authors carried out the procedures for this project including study design, execution, and acquisition of data, analysis and interpretation. GUE supervised the overall conceptual design, implementation of the project, and revision of the manuscript. All authors agreed on the journal to which the article was submitted. All authors was involved in the writing and substantial revision of this manuscript. All authors contributed to the critical review and final approval of the manuscript. All the authors read, approved the final manuscript and agreed to be accountable for all aspects of the work.

## Funding

The research was funded by the Mojeaga International Ventures Ltd, Nigeria and the researchers. The funders had no role in the design, conduct, analysis, interpretation, or write up of the study.

## Data sharing statement

All relevant data are within the manuscript and its Supporting Information files. The datasets used and/or analyzed during the current study are available from the authors on reasonable request.

## Ethical approval and informed consent

The study protocol was approved by the Nnamdi Azikiwe University Teaching Hospital, Nnewi, Nigeria ethics committee and other collaborating hospitals, with the approval numbers: NAUTH/CS/66/VOL.12/014/2019/008, ESUTHP/C-MAC/RA/034/103, and COOUTH/CMAC/ETH.C/VOL.1/0056. The study was also prospectively registered at www.pactr.samrc.ac.za (with registered protocol available at: https://pactr.samrc.ac.za/TrialDisplay.aspx?TrialID=5822) with a registration number of PACTR201901852059636. The trial was also registered, approved and monitored by National agency for food drug and control (NAFDAC) available at https://www.nafdac.gov.ng/ with NAFDAC Trial Registration No of NAF/DER/LAG/V&CT/MOJEAGA/2021. The objectives and purposes of the studies, as well as risks and benefits was explained to the participants. Participants were informed that participation in the study is voluntary; they have the right to withdraw or refuse to participate in the study at any time. Written and signed informed consent were obtained from each study participant prior to interview as it is stated in participant information sheet. The data collection procedure was anonymous in order to keep the confidentiality of any information provided by the study participants.

## Consent for publication

Not applicable.

## Competing interests

The authors declare that they have no competing interests. The abstract of this preliminary data was presented at the virtual XXIII World Congress of Gynecology and Obstetrics, Australia, in October 2021.

## Notes

### Competing Interest Statement

The authors have declared no competing interest.

### Clinical Trial

Clinical Trial Registration: www.pactr.samrc.ac.za: PACTR201901852059636.

### Funding Statement

This study was (partially) funded by the Mojeaga International Ventures Ltd, Nigeria. The funders had no role in study design, data collection and analysis, decision to publish, or preparation of the manuscript.

### Author Declarations

The study protocol was approved by the Nnamdi Azikiwe University Teaching Hospital, Nnewi, Nigeria ethics committee and other collaborating hospitals, with the approval numbers: NAUTH/CS/66/VOL.12/014/2019/008, ESUTHP/C-MAC/RA/034/103, and COOUTH/CMAC/ETH.C/VOL.1/0056.

